# Improvement of Survival Outcomes of Cholangiocarcinoma by Ultrasonography Surveillance: Multicenter Retrospective Cohorts

**DOI:** 10.1101/2024.02.29.24303543

**Authors:** Nittaya Chamadol, Vallop Laopaiboon, Apiwat Jareanrat, Vasin Thanasukarn, Tharatip Srisuk, Vor Luvira, Poowanai Sarkhampee, Winai Ungpinitpong, Phummarat Khamvijite, Yutthapong Chumnanua, Nipath Nethuwakul, Passakorn Sodarat, Samrit Thammarit, Anchalee Techasen, Jaruwan Thuanman, Chaiwat Tawarungruang, Bandit Thinkhamrop, Prakasit Sa-Ngiamwibool, Watcharin Loilome, Piya Prajumwongs, Attapol Titapun

**Author notes:** **Corresponding author:** (AT).

## Abstract

**Introduction:** Most cholangiocarcinoma (CCA) patients present with late stage of disease because of the difficulty to diagnosis at an early stage, resulting in poor survival of CCA patients. The Cholangiocarcinoma Screening and Care Program showed that ultrasound screening was an effective tool for detecting early stage CCA. This study aims to evaluate the survival outcome of patients diagnosed by ultrasound screening (US) compared to walk-in symptomatic patients.

**Methods:** 5-year survival rates (5-YSR) and median survival time (MST) of CCA were calculated using Log-Rank test. Multivariate analyses were performed for significant factors from univariate analyses.

**Results:** A total of 711 histologically proven CCA cases were examined including ultrasound screening and walk-in groups. The screening group having 5-YSR was 53.9%, and MST was of 67.2 months, while walk-in group, the 5-YSR was 21.9% and MST was 15.6 months (*p*<0.001). In addition, multivariate analyses revealed that screening program was an independent factor to predict a good outcome of CCA patients when compared with walk-in group (*p* = 0.014).

**Conclusion:** US is an effective tool for detecting early stage CCA leading to improve clinical outcome of CCA patients. Practically, US should be considered as a first tool for screening CCA in risk populations.

**Author Summary:** Most cholangiocarcinoma (CCA) patients in Thailand have poor survival due to late-stage detection and patients walk-in to hospital with any symptoms. This study purpose to evaluate the survival outcome of CCA patients diagnosed by ultrasound screening (US). We found that US provided early stage and improved survival of CCA patients.

## Introduction

Cholangiocarcinoma (CCA) is a cancer of bile duct epithelium which the second most common primary liver cancer worldwide after hepatocellular carcinoma (HCC). CCA has a relatively rare incidence in most western countries and North America, however, it has been reported as having high incidence rates in East and Southeast Asia, especially in Thailand. The incidence of CCA in the northeast of Thailand has been recorded as the highest incidence rate worldwide with an incidence of 87.7 per 100,000 in males, and 36.3 per 100,000 in females [1]. The major risk factor of CCA in Thailand has been identified as infection by the liver fluke *Opisthorchis viverrini* (OV) which initiates the development of the normal bile duct to transform into tumor known as cholangiocarcinogenesis [2–4].

CCA patients have poor survival and high mortality rate due to late diagnosis. Diagnosis of CCA is rare at an early stage because most patients with any clinical symptoms were diagnosed at the advance or locally advance stage. Surgical resection is potentially the most curative treatment considered as a first choice for treatment in resectable patients in every type of CCA [5, 6]. Surgical resection offers the best opportunity for long-term survival with survival time approximately 17-20 months and 5-year survival rate 10-25% [7–12]. Although surgical resection provides long-term overall survival, candidate surgical patients have been reported to be only 20%, while 80% are diagnosed at unresectable stage CCA [13]. The unresectable CCA patients suffer from several complications for instance, local tumor invasion or distant metastasis, biliary obstruction, cholangitis, pain, and malnutrition [13]. These complications reduced the quality of the patient’s life, with subsequent poor survival of unresectable CCA patients. Thus, a screening test for diagnosis of CCA at an early stage of disease has diagnostic and clinical advantages for the early treatment of CCA which improves a patient’s outcomes.

Trans-abdominal ultrasonography (US) is a non-invasive imaging tool to detect abnormality in the hepatobiliary system, including early stage CCA by detecting mass and/or dilatation of bile ducts. US also offers several advantages, due to its accessibility, speed, ease of performance, portability and low cost [14]. Therefore, US should be considered as the first-choice imaging modality for screening abnormalities associated with CCA in Thailand [15–19]. Ultrasound screening was systematically applied in the Cholangiocarcinoma Screening and Care Program (CASCAP), to determine the utility of the application for early diagnosis of CCA combined with prevention, treatment, and follow-up. This prospective study consisted of two cohorts, the screening cohort included people at risk of CCA without any symptoms who received active ultrasound screening, and the patient cohort included symptomatic walk-in patients [20]. Results showed that US screening can diagnose early signs of biliary tract fibrosis (periductal fibrosis) that is associated with CCA [15] as well as detecting premalignant CCA lesions and early stage CCA [21]. Subsequently, CCA patients who were diagnosed by US screening had significantly higher proportion of early stage CCA compared to symptomatic walk-in patients [17]. Early-stage detection in the screening group may provide better survival outcomes than the walk-in group of CCA, significant benefits for early treatment and reduction of morbidity and mortality rates of CCA patients.

Therefore, this study aimed to evaluate the effectiveness of US screening by comparing the survival outcome between the screening group and symptomatic walk-in patient group.

## Methods

### Ethics statement and consent to participate

This study was conducted based on the principles of Good Clinical Practice, the Declaration of Helsinki, and national laws and regulations about clinical studies. All processes of this study were accepted and approved by the Khon Kaen University Ethics Committee for Human Research under the reference number HE551404. The data underwent complete anonymization before our access. Information could not identify individual participants. Clinical data and medical records of patients were retrieved through only hospital number (HN). Clinical data and medical records for this study were accessed in 30 June 2021.

### Overview of study design

A total of 766 CCA patients were included in this study who underwent surgery and CCA was confirmed by pathologists in 11 hospitals over the period of 1 October 2013 to 30 June 2021, namely: (1) Srinagarind Hospital, Khon Kaen University; (2) Sunpasittiprasong Hospital; (3) Surin Hospital; (4) Udon Thani Hospital; (5) Roi Et Hospital; (6) Udonthani Cancer Hospital; (7) Maharaj Nakorn Chiang Mai Hospital; (8) Buri Ram Hospital; (9) Ubonratchathani Cancer Center; (10) Khon Kaen Hospital and (11) Maharat Nakhon Ratchasima Hospital. Demographic and pathological data that were recorded by the Cholangiocarcinoma Research Institute (CARI), Khon Kaen University were reviewed. Patients in this study were separated into two groups: the screening group (n = 163) comprised individuals diagnosed by US screening who had no clinical symptoms that could be related to CCA and the walk-in group (n = 603) which comprised patients presenting with clinical symptoms and confirmed CCA by CT/MRI. A total of 55 patients were excluded from the study as they did not receive curative surgery due to the advanced CCA stage, unresectable or had distant metastasis (5 cases in screening and 23 cases in walk in groups), and patients who had survival time less than 30 days (4 cases in screening and 23 cases in walk in groups). Therefore, a total of 711 cases were included in this study comprising the screening group of 154 cases and walk-in group of 557 cases (Fig 1).

**Fig 1.**
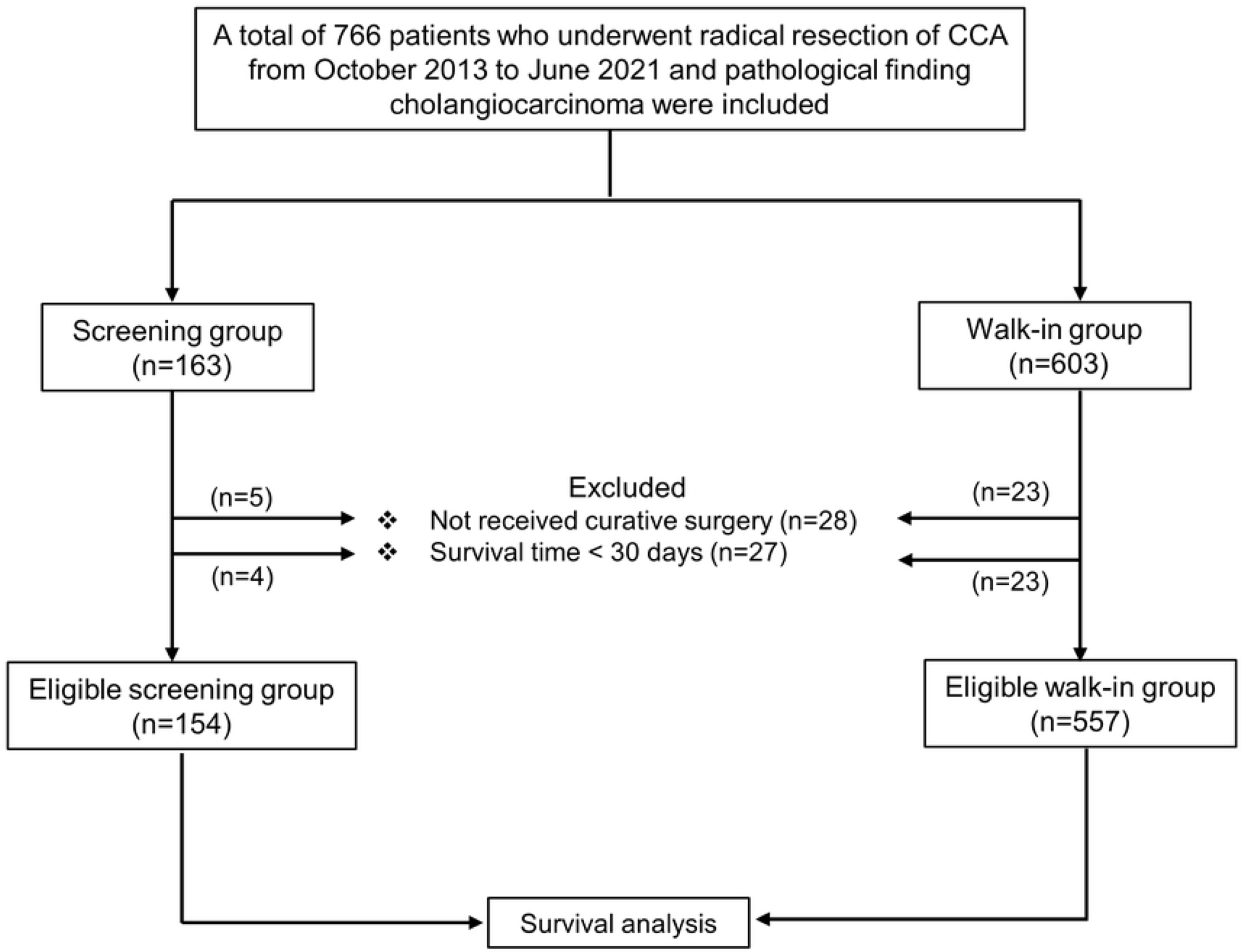
The schematic of the study.

### Diagnosis and treatment

Patients in screening groups who underwent abdominal US examination and confirmed by CT/MRI. US and CT/MRI images of both groups were reviewed by radiologist (NC and VL). Intraoperative findings and operative procedure were reviewed. Histopathological diagnosis and tumor morphology of both groups were reviewed by pathologists (PS). Tumor staging was recorded according to AJCC 8^th^ edition. Adjuvant chemotherapy was provided to patients by attended oncologists or by a multidisciplinary team conference at each treatment center. Patients were followed up with CT/MRI and tumor markers every 3-6 months. If recurrence of disease occurred, a different chemotherapy regimen was considered and applied for appropriate patients.

### Data management and statistical analysis

The demographic characteristics of the patients were presented as the mean and standard deviation for continuous variables and frequency counts with their percentages for categorical variables. Both of these were presented for each comparison group and as a total for the purpose of comparison and characterizing the patient cohort, respectively. The proportion of early stage CCA was calculated by using the number of patients whose stage was 0, I, or II as the numerator and the total number of patients as the denominator.

Survival analysis was calculated by Kaplan-Meier method. Survival time was defined as from the date of surgery to the date of the patient’s death. Patients who survived after the end of study date (30^th^ December 2021) were defined as censor. Median survival times and survival rates are presented with 95%CI and the comparison between groups was analyzed by log rank test. Univariable and multivariable analyses were performed to identify prognostic factors using the Cox regression model. A *p*-value of less than 0.05 was considered significant. All analyses were performed using the IBM SPSS Statistics version 26.

## Results

### Patient’s characteristics and overall survival of CCA patients between screening and walk-in

The number and proportion of CCA patients who received curative surgery in 11 hospitals are as follows: (1) Srinagarind Hospital, Khon Kaen University, n = 493 (69.3%); (2) Sunpasittiprasong Hospital, n = 62 (8.7%); (3) Surin Hospital, n = 48 (6.7%), (4) Udon Thani Hospital, n = 44 (6.2%); (5) Roi Et Hospital, n = 41 (5.8%); (6) Udonthani Cancer Hospital, n = 8 (1.1%); (7) Maharaj Nakorn Chiang Mai Hospital, n = 7 (1%); (8) Buri Ram Hospital, n = 4 (0.6%); (9) Ubonratchathani Cancer Center, n = 2 (0.3%); (10) Khon Kaen Hospital, n = 1 (0.15%) and (11) Maharat Nakhon Ratchasima Hospital, n = 1 (0.15%).

A total of 711 cases of CCA patients were included in this study and separated into 2 groups namely, 154 (21.7%) cases for the screening group and 557 (78.3%) cases for the walk-in group. The median age of patients was 61 years, where the majority were found to be male 451 (63.4%). Intrahepatic CCA (iCCA) 349 (49.3%) cases were found to be the highest in this study, followed by perihilar (pCCA) and distal (dCCA) CCA 282 (39.9%) and 76 (10.8%) cases, respectively. The screening groups had significantly higher proportions of iCCA than the walk-in groups, while pCCA and dCCA were found to be significantly greater in the walk-in group (*p* < 0.001). Results of tumor morphology showed that the mass-forming types were the major subtypes 300 (43.0%) cases in both groups. For tumor staging according to the AJCC/UICC staging system, tumor stage was categorized into two groups, early stage (0-II), 254 (35.7%), and late stage (III-IV), 457 (64.3%). Interestingly, tumor staging was separated based on programs to detect CCA. Result showed that screening groups had significantly higher CCA patients with early stage CCA 130/154 (84.4%) than walk-in groups with 124/557 (22.3%) cases (*p* < 0.001) (Supplementary Table 1).

The survival analysis was performed to calculate 5-year survival rate (5-YSR) and median survival time (MST) presenting by month. The overall survival of 711 patients of this study showed that MST was 19.9 months, and 5-YSR was 28.8% (Supplementary Fig 1). Age, gender, and anatomical locations had no significant effect on the 5-YSR and MST in this study. However, tumor morphology showed that patients with ID had significantly better survival than PI, MF, and mixed type (5-YSR = 47.7 *vs.* 27.1, 24.0 and 22.9%; MST = 44.3 *vs.* 20.2, 14.3 and 23.7 months; HR = 1.62, 1.89 and 1.66, *p* = 0.001, < 0.001 and 0.012, respectively). The comparison of the survival in early and late stage showed that patients with early stage had markedly greater survival than patients with late stage (5-YSR = 54.7 *vs.* 14.4%; MST = 78.4 *vs.* 12.3 months; HR = 3.40, *p* < 0.001). Interestingly, patients in the walk in group had significantly greater 5-YSR and MST than the walk in group (5-YSR = 53.9 *vs.* 21.9%; MST = 67.2 *vs.* 15.6 months; HR = 2.61, *p* <0.001) (Table 1 and Fig 2).

**Fig 2.**
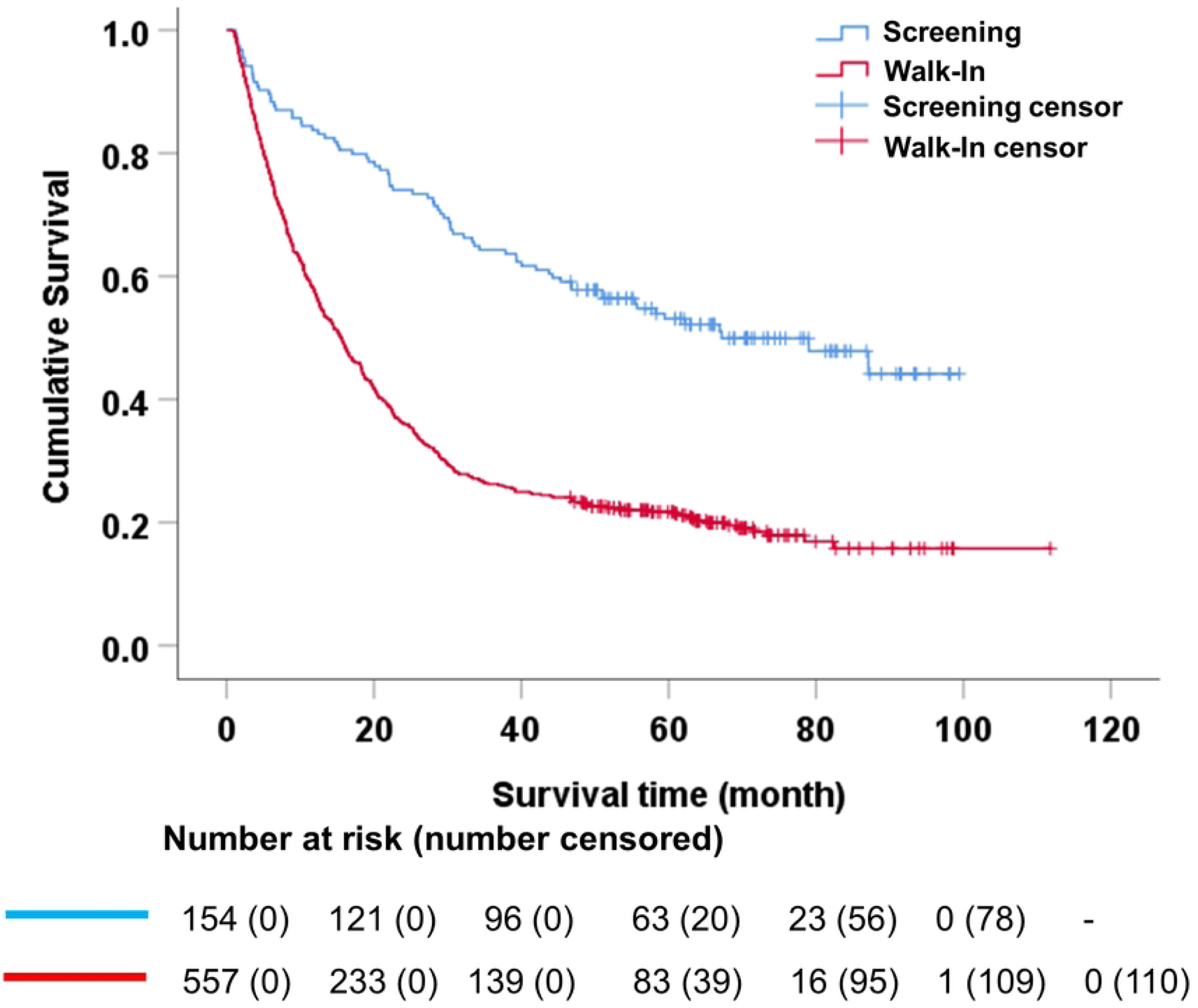
The survival of CCA patients in screening and walk-in groups.

**Table 1.** Univariate and multivariate analysis of the survival of CCA patients.

| Variable | n | 5-YSR (%)<br>(95%CI) | MST (month)<br>(95%CI) | Univariate |  | Multivariate |  |
| --- | --- | --- | --- | --- | --- | --- | --- |
|  |  |  |  | HR<br>(95%CI) | *p*-value | HR<br>(95%CI) | *p*-value |
| Age, years |  |  |  |  |  |  |  |
| < 61 | 326 | 26.4<br>(22.2-30.6) | 19.0<br>(15.5-22.5) | 1 |  | - |  |
| ≥ 61 | 385 | 30.9<br>(26.7-35.1) | 20.3<br>(16.1-24.4) | 0.93<br>(0.78-1.09) | 0.378 | - | - |
| Gender |  |  |  |  |  |  |  |
| Male | 451 | 29.7<br>(25.9-33.5) | 20.1<br>(17.1-23.1) | 1 |  | - |  |
| Female | 260 | 27.3<br>(22.5-32.1) | 18.4<br>(13.9-22.8) | 0.95<br>(0.79-1.13) | 0.542 | - | - |
| Tumor<br>Location<sup>a</sup> |  |  |  |  |  |  |  |
| dCCA | 76 | 36.8<br>(26.5-47.1) | 22.4<br>(18.7-26.1) | 1 |  | - |  |
| iCCA | 349 | 30.4<br>(26.1-34.7) | 19.6<br>(15.6-23.6) | 1.23<br>(0.92-1.71) | 0.147 | - | - |
| pCCA | 282 | 25.2<br>(20.8-29.6) | 18.9<br>(15.7-22.0) | 1.36<br>(0.99-1.86) | 0.054 | - | - |
| Tumor<br>Morphology<sup>a</sup> |  |  |  |  |  |  |  |
| ID | 128 | 47.7<br>(39.4-56.0) | 44.3<br>(17.9-70.7) | 1 |  | 1 |  |
| PI | 221 | 27.1<br>(21.7-32.5) | 20.2<br>(16.4-24.1) | 1.62<br>(1.45-2.45) | 0.001* | 1.25<br>(0.94-1.65) | 0.126 |
| MF | 300 | 24.0<br>(19.9-28.1) | 14.3<br>(10.8-17.8) | 1.89<br>(1.23-2.13) | < 0.001* | 1.36<br>(1.04-1.78) | 0.025* |
| Mix | 48 | 22.9<br>(15.0-30.8) | 23.7<br>(11.5-35.8) | 1.66<br>(1.12-2.47) | 0.012* | 0.96<br>(0.64-1.44) | 0.847 |
| TNM stage |  |  |  |  |  |  |  |
| Early (0-II) | 254 | 54.7 | 78.4 | 1 |  | 1 |  |
|  |  | (48.6-60.8) | (59.7-97.2) |  |  |  |  |
| Late (III-IV) | 457 | 14.4<br>(11.8-16.9) | 12.3<br>(10.5-14.1) | 3.40<br>(2.76-4.18) | < 0.001* | 2.76<br>(2.15-3.55) | < 0.001* |
| Diagnostic methods |  |  |  |  |  |  |  |
| Screening | 154 | 53.9<br>(46.1-61.7) | 67.2<br>(44.1-90.2) | 1 |  | 1 |  |
| Walk-In | 557 | 21.9<br>(18.9-24.9) | 15.6<br>(13.4-17.9) | 2.61<br>(2.05-3.34) | < 0.001* | 1.44<br>(1.10-1.92) | 0.014* |

n, Number; CI, Confidence interval; 5-YSR, 5-year survival rate; MST, median survival time; HR, hazard ratio; dCCA, distal cholangiocarcinoma; iCCA, Intrahepatic cholangiocarcinoma; pCCA, perihilar cholangiocarcinoma; ID, intraductal; PI, periductal infiltrating; MF, mass-forming; TNM, tumor node metastasis from 8<sup>th</sup> AJCC/UICC staging system.
<sup>a</sup> The data was not available in some case
\* Indicates a *p*-value < 0.05 (statically significant)

The significant factors of the survival determined by univariate investigations were further analyzed to identify any independent factor(s) for use as prognostic prediction of the outcome of CCA patients which was composed of tumor morphology, staging and diagnostic methods. The multivariate analysis showed that MF morphology, late CCA stage and the walk-in group were statistically independent factors for poor prognosis (HR = 1.36, *p* = 0.025; HR = 2.76, *p* < 0.001; and HR = 1.44, *p* = 0.014, respectively) (Table 1).

### Subgroup analysis of screening and walk-in groups on the survival outcomes of CCA patients

Subgroup analysis of each variable in both screening and walk-in groups showed no difference in the survival outcome by age and gender.

Tumor morphology comparisons showed ID had better survival than PI, MF, and mixed type in both the screening and walk in group. Early stage of disease was factor in good survival of patients in both groups. There was a different outcome in tumor location, where results showed that iCCA and pCCA had a good 5-YSR than dCCA (52.4 and 65.9 *vs.* 22.2%, *p* < 0.05, respectively). In contrast, the in walk in group, dCCA had better 5-YSR than iCCA and pCCA (38.8 *vs.* 21.1 and 18.3%, *p* < 0.001, respectively) (Table 2 and Fig 3).

**Fig 3.**
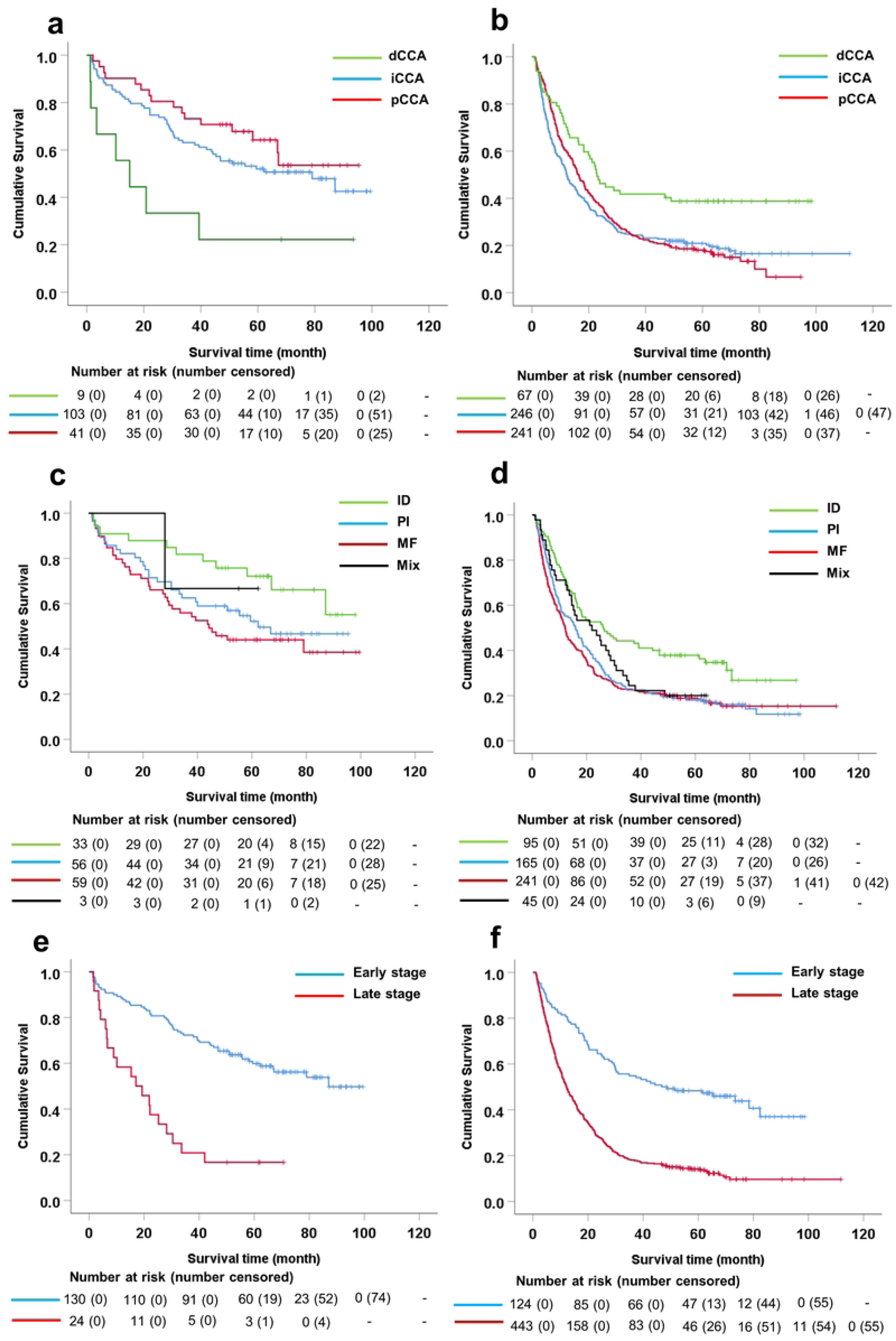
Subgroup analysis of the survival in screening and walk in group. (A) Survival curve of tumor location in screening and (B) walk in, (C) tumor morphology in screening and (D) walk in and (E) tumor staging in screening and (F) walk in.

**Table 2.**
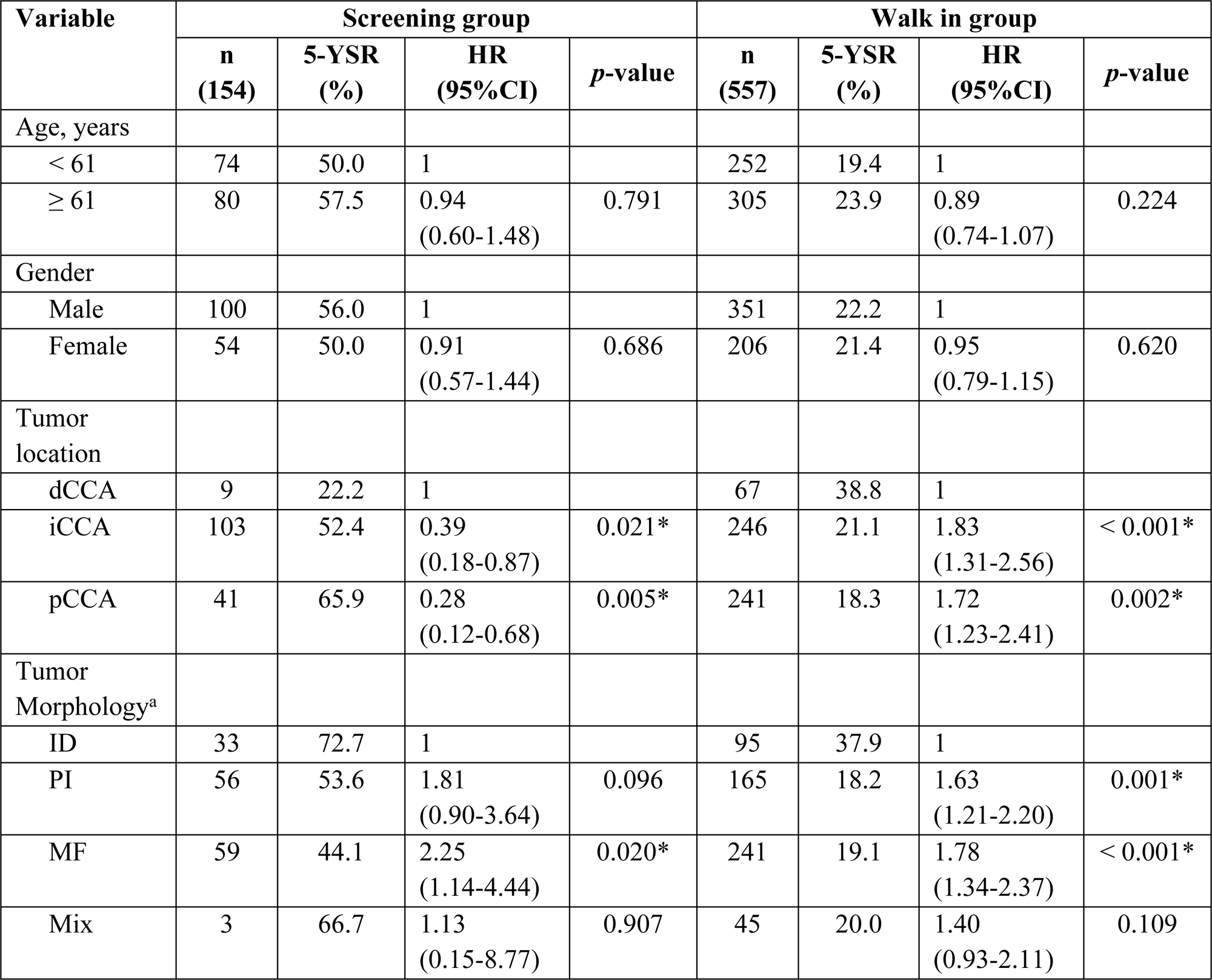
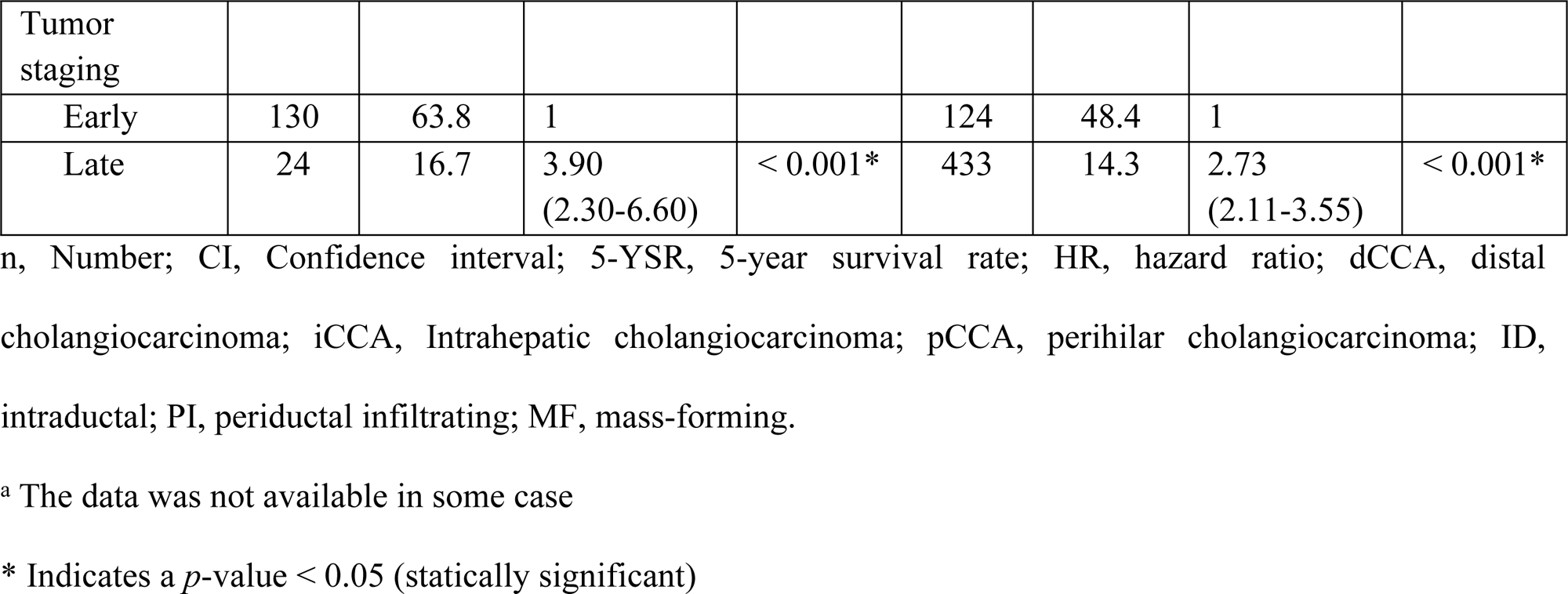
The comparison of the survival in CCA patients between screening and walk in methods.

| Variable | Screening group |  |  |  | Walk in group |  |  |  |
| --- | --- | --- | --- | --- | --- | --- | --- | --- |
|  | n<br>(154) | 5-YSR<br>(%) | HR<br>(95%CI) | *p*-value | n<br>(557) | 5-YSR<br>(%) | HR<br>(95%CI) | *p*-value |
| Age, years |  |  |  |  |  |  |  |  |
| < 61 | 74 | 50.0 | 1 |  | 252 | 19.4 | 1 |  |
| ≥ 61 | 80 | 57.5 | 0.94<br>(0.60-1.48) | 0.791 | 305 | 23.9 | 0.89<br>(0.74-1.07) | 0.224 |
| Gender |  |  |  |  |  |  |  |  |
| Male | 100 | 56.0 | 1 |  | 351 | 22.2 | 1 |  |
| Female | 54 | 50.0 | 0.91<br>(0.57-1.44) | 0.686 | 206 | 21.4 | 0.95<br>(0.79-1.15) | 0.620 |
| Tumor location |  |  |  |  |  |  |  |  |
| dCCA | 9 | 22.2 | 1 |  | 67 | 38.8 | 1 |  |
| iCCA | 103 | 52.4 | 0.39<br>(0.18-0.87) | 0.021* | 246 | 21.1 | 1.83<br>(1.31-2.56) | < 0.001* |
| pCCA | 41 | 65.9 | 0.28<br>(0.12-0.68) | 0.005* | 241 | 18.3 | 1.72<br>(1.23-2.41) | 0.002* |
| Tumor Morphology<sup>a</sup> |  |  |  |  |  |  |  |  |
| ID | 33 | 72.7 | 1 |  | 95 | 37.9 | 1 |  |
| PI | 56 | 53.6 | 1.81<br>(0.90-3.64) | 0.096 | 165 | 18.2 | 1.63<br>(1.21-2.20) | 0.001* |
| MF | 59 | 44.1 | 2.25<br>(1.14-4.44) | 0.020* | 241 | 19.1 | 1.78<br>(1.34-2.37) | < 0.001* |
| Mix | 3 | 66.7 | 1.13<br>(0.15-8.77) | 0.907 | 45 | 20.0 | 1.40<br>(0.93-2.11) | 0.109 |

| Tumor staging |  |  |  |  |  |  |  |  |
| --- | --- | --- | --- | --- | --- | --- | --- | --- |
| Early | 130 | 63.8 | 1 |  | 124 | 48.4 | 1 |  |
| Late | 24 | 16.7 | 3.90<br>(2.30-6.60) | < 0.001* | 433 | 14.3 | 2.73<br>(2.11-3.55) | < 0.001* |

n, Number; CI, Confidence interval; 5-YSR, 5-year survival rate; HR, hazard ratio; dCCA, distal cholangiocarcinoma; iCCA, Intrahepatic cholangiocarcinoma; pCCA, perihilar cholangiocarcinoma; ID, intraductal; PI, periductal infiltrating; MF, mass-forming.
<sup>a</sup> The data was not available in some case
\* Indicates a *p*-value < 0.05 (statically significant)

## Discussion

Cholangiocarcinoma (CCA) is most common primary malignancy of bile duct epithelia in the biliary tract. The northeast of Thailand has the highest incidence of CCA in the world [22]. Most CCA patients are diagnosed at a late stage of the disease leading to poor survival of patients due to cancer metastasis. Studies have shown that approximately 20,000 CCA patients die in northeast Thailand per year leading to a significant socioeconomic burden for the affected families [23]. Therefore, early diagnosis is important to enable appropriate early treatment plan to be implemented, and hence, improve patient outcomes.

A previous report by Luvira V *et al* showed that only 20% of CCA patients were treated with surgical resection, while 80% of CCA patients were unresectable cases who had palliative treatments such as symptomatic treatment, chemotherapy, palliative drainage and biliary stent insertion [13]. Unfortunately, despite these palliative measures overall survival is still poor due to the advanced stage of disease leading to cancer metastasis [13, 24]. In addition, several confounding complications and symptoms present at late stage, such as biliary obstruction, obstructive jaundice and cholangitis which reduce quality of life of patients [13, 25–27]. Our study showed that patients with late stage CCA was still currently high at approximately 64.3% while patients with early stage CCA was 35.7%. The overall survival rate and median survival time of CCA patients after curative surgery was 28.8% and 18.5 months which was concordant with the range of survival outcome of CCA patients in previous reports of approximately 10-25% and 17-20 months, respectively [7–12].

In 2020, Khuntikeo *et al* evaluated the efficiency of different methods for CCA detection by comparing of screening programs using US by the CASCAP program (screening group) and participant walk-in with clinical symptom hospital group in 762 histologically proven CCA cases. Results showed that the proportion of early stage CCA in the screening group (0-II) was 84.5%, while it was 21.6% in walk-in group. The comparison suggested that US via active screening improves early-stage detection and was significantly higher than the walk-in group, hence US is an effective tool for detecting early-stage disease of CCA [17]. The present study was a retrospective study incorporating 11 hospitals in Thailand to compare monitoring methods comprising the screening group and a walk-in group. The 11 hospitals conducted consensus procedures for US screening the suspected patients in high-risk areas, while patients who came to hospital with any symptoms were classified into the walk-in group. Subsequently, all patients in both groups were enroll for curative treatment by surgical resection, and comparison of the survival outcomes of CCA between the screening group and a walk-in group was undertaken. The results showed that screening groups provided significantly better survival rate and median survival than the walk-in groups (53.9 vs 21.9% and 67.2 vs 15.6 months, respectively (Table 1). This good survival outcome was concordant with the higher proportion of early stage CCA in the screening groups (84.4%) than the walk-in groups (22.3%) by around 4-fold (Supplementary Table 1). Moreover, the walk-in groups had more patients excluded from study due to advanced stage and received only palliative surgery with mortality within 30 days. This finding may reflect advanced CCA staging and risk of surgery in the walk-in group compared to screening group.

The screening group provides a greater number of early stage CCA patients than walk-in group because in the US-screening program by the CASCAP, suspected CCA cases without symptoms in the high risk CCA endemic area are screened early by US, therefore, premalignant lesions or early stage is usually found at the earliest possible detection time. Conversely, in the walk-in group, patients present to hospital with abnormality or clinical symptoms. These clinical symptoms are frequently correlated with CCA at an advanced stage and recorded as abnormal at the hospital admission [17, 25, 27, 28]. Therefore, results from our study show that early screening is most important to enable detection of early stage of the disease which can then provide appropriate early treatment and surveillance of patients to improve their overall survival outcome.

In addition, a significant finding of this study was survival outcomes concerning tumor location which was obviously different. For the screening group, patients with iCCA and pCCA had longer survival times than dCCA of approximately 2-fold, while in walk in groups, we found that patients with dCCA had better survival than iCCA and pCCA of around 2-fold. These results can be explained because US has an effective capability for detection of early lesion in liver parenchyma, such as small nodules, periductal fibrosis, focal duct dilatation, however, but some limitation of US to detect distal bile duct lesions was observed. Distal bile duct tumor can be detected by US by detecting common bile duct dilatation while most of patients with common bile duct dilatation were not an early stage or had obstructive symptom already.

According to several publications, most CCA patients in Thailand were iCCA and pCCA, and they come to hospital with symptoms which are diagnosed as late or advance stage of disease, such as severe extension, lymph node and distant metastasis [29–31]. Our results also showed that generally 64.3% of patients present at late stage disease. In contrast, although dCCA is also present at late stage, it causes a symptom more readily. Therefore, a 5-YSR for patients with dCCA (38.8%) was significantly better than iCCA (21.1%) and pCCA (18.3%) as presented in walk in groups (Table 2). US screening has been reported as a tool for the early detection of premalignant lesions and early stage of CCA [15–21]. The suspected CCA cases without symptoms from preliminary detection were diagnosed as all types of CCA, especially iCCA and pCCA for which there are no symptoms until advanced stage, resulting in early treatment, surveillance, and improvement of overall survival. Our results showed that a 5-YSR of patients with iCCA and pCCA was markedly better than patients in walk in group. Furthermore, since early monitoring was performed in screening group, almost all of the CCA patients, especially those with iCCA and pCCA, had good survival than patients in the walk-in group, which leads to improved effective treatment, surveillance, and survival outcome in CCA patients.

Additionally, we found that patients with early stage in screening group had 5-YSR better than walk in group (63.8 vs 48.4%, respectively) and received early management and treatment plan. Conversantly, patients having early stage in the walk-in group have some symptoms such as sepsis, malnutrition, poor physical status [17, 25, 27, 28]. These symptoms may result in poorer outcomes despite patients being in the early stage of disease. However, in the late stage of disease there were no differences in the survival outcome of both groups as several independent factors can affect on the survival of patients.

Tumor staging is well known to have an affect on the patient’ survival outcome. Tumor morphology is also a potential factor to predict the survival outcome of CCA patients. Tumor morphology has been classified into four types, mass-forming (MF), periductal-infiltrating (PI), intraductal (ID), and mixed types. Basically, ID is represented as good survival in tumor morphology while PI and MF are associated with aggressive features and poor survival of patients [12, 21, 32–38]. This information has recently been confirmed and shows that tumor morphology relates and predicts the survival outcome of all types of CCA after curative surgical treatment. Results from our study also showed that tumor morphology could be a predictor of survival outcomes. For instance, ID was obviously associated with longer survival than PI and MF. Moreover, subgroups analysis of tumor morphology for patient’s survival of screening and walk-in groups also showed a similar outcome of ID having better survival than PI and MF. This result could explain that screening programs had no effect on changing biology of tumor morphology to impact on CCA patient’s survival. Nevertheless, results of the screening group highlight that all types of tumor morphology had markedly better 5-YSR than those in walk-in group.

Although this study showed several advantages of US in suspected cases who may be CCA, there is some limitation in an imbalance of numbers and variables between screening and walk in groups.

In summary, this finding revealed that ultrasound screening for CCA is an effective tool for detecting early stage CCA, and significantly improves survival outcome of CCA patients. Therefore, a comprehensive population-based programs using US for screening early stage CCA in areas of high incidence throughout Thailand and elsewhere in Southeast Asia should be undertaken.

## Data Availability

All relevant data are within the manuscript and its Supporting Information files.

## Acknowledgements

All authors are truly thankful Prof. Narong Khuntikeo at Department of Surgery, Faculty of Medicine, Khon Kaen University, Khon Kaen, Thailand, Cholangiocarcinoma Research Institute (CARI), Khon Kaen University, Khon Kaen, Thailand and Cholangiocarcinoma Screening and Care Program (KKU), Khon Kaen University, Khon Kaen, Thailand for helpful discussions. We are also indebted to all members of CASCAP, particularly the cohort members, and researcher at CARI, Faculty of Medicine, Khon Kaen University for collecting and proofing of CCA patient data. In addition, we also thank Professor Ross H. Andrew for editing the MS via the Publication Clinic KKU, Thailand.

## Author Contributions

**Conceptualization:** Nittaya Chamadol, Watcharin Loilome, Attapol Titapun.

**Funding acquisition:** Nittaya Chamadol, Watcharin Loilome, Attapol Titapun.

**Sample collection and diagnosis**: Nittaya Chamadol, Vallop Laopaiboon, Apiwat Jareanrat, Vasin Thanasukarn, Tharatip Srisuk, Vor Luvira, Poowanai Sarkhampee, Winai Ungpinitpong, Phummarat Khamvijite, Yutthapong Chumnanua, Nipath Nethuwakul, Passakorn Sodarat, Samrit Thammarit, Prakasit Sa-Ngiamwibool, Attapol Titapun.

**Analysis and interpretation of data:** Jaruwan Thuanman, Chaiwat Tawarungruang, Bandit Thinkhamrop, Piya Prajumwongs, Attapol Titapun.

**Project administration:** Nittaya Chamadol, Watcharin Loilome, Attapol Titapun.

**Supervision:** Nittaya Chamadol, Watcharin Loilome, Attapol Titapun.

**Validation:** Nittaya Chamadol, Vallop Laopaiboon, Apiwat Jareanrat, Vasin Thanasukarn, Tharatip Srisuk, Vor Luvira, Poowanai Sarkhampee, Winai Ungpinitpong, Phummarat Khamvijite, Yutthapong Chumnanua, Nipath Nethuwakul, Passakorn Sodarat, Samrit Thammarit, Anchalee Techasen, Jaruwan Thuanman, Chaiwat Tawarungruang, Bandit Thinkhamrop, Prakasit Sa-Ngiamwibool, Watcharin Loilome, Piya Prajumwongs, Attapol Titapun.

**Writing original draft:** Nittaya Chamadol, Watcharin Loilome, Piya Prajumwongs.

**Writing review and editing:** Nittaya Chamadol, Anchalee Techasen, Watcharin Loilome, Piya Prajumwongs, Attapol Titapun. All authors approved the final version of the manuscript.

## Supporting information captions

**S1 Table. Patient characteristics.** n−Number; CI−Confidence interval; 5-YSR−5-year survival rate; dCCA−distal cholangiocarcinoma; iCCA−Intrahepatic cholangiocarcinoma; pCCA−perihilar cholangiocarcinoma; ID−intraductal; PI−periductal infiltrating; MF−mass-forming. ^a^ The data was not available in some case. ^$^ Indicates the data were testes by Fisher’s exact test. * Indicates a *p*-value < 0.05 (statically significant).

**S1 Fig. Overall survival of CCA patients in the study.**

